# Exploring the use of synthetic placebo populations in ALS randomized clinical trials

**DOI:** 10.1101/2022.12.09.22283281

**Authors:** Harry Bowles, Sarah Opie-Martin, Ali Shojaie, Laura Libonati, Alfredo Iacoangeli, Ahmad Al Khleifat, Ammar Al-Chalabi

**Author notes:** both authors contributed equally to this work (Harry Bowles, Sarah Opie-Martin). Harry Bowles is a PhD student working on bioinformatics and clinical trial optimisation in amyotrophic lateral sclerosis. His research is focused on genome sequence analysis and synthetic data. Dr Sarah Opie-Martin is an epidemiologist working on data collection and analysis of epidemiology projects in amyotrophic lateral sclerosis. Her research focusses on environmental risk factors for ALS, clinical subgroups, and disease prognosis. Ali Shojaie is a medical doctor and clinical researcher. He is doing his PhD in clinical neuroscience. His PhD focused on different aspects of non-motor symptoms in ALS. Laura Libonati is a PhD student working in the clinical setting with patients with Amyotrophic Lateral Sclerosis. Her research is focused on analyzing clinical, electrophysiological, and serological markers with a possible role in the disease’s development and progression. After a career in Biophysics, Dr. Alfredo Iacoangeli accepted his PhD “cum laude” in Life Sciences in January 2016 from “Sapienza” University of Rome where he worked on structural bioinformatics with a particular focus on protein structure prediction and protein-peptide interaction. He joined King’s College London in March 2016 and he is now the Bioinformatics lead of a joint programme between the Department of Basic and Clinical Neuroscience and the Health Informatics Unit at the Institute of Psychiatry, Psychology & Neuroscience at King’s College London. The aim of this project is twofold: 1) the development of a high throughput gene, environment and epigenetics database and analysis system for international MND/ALS research; 2) the use of large multi-omics datasets to identify subgroups MND/ALS patients with homogeneous disease causes and clinical phenotype, and to gain new insights into the disease pathogenesis. Dr. Iacoangeli is the primary author of several scientific articles in the field of Structural and Genomic Bioinformatics and MND/ALS genetics. Dr Ahmad Al Khleifat is a clinician and scientist currently working with Professor Al-Chalabi’s group. His main focus of research is on ALS clinical staging, clinical trial data analysis, and disease gene identification through next generation sequencing and coupling this with advanced data analysis to deliver diagnostic tools for complex disease genetics. Since 2017 Dr Al Khleifat has been a co-chair of genomic structural variations group responsible for the collection of the largest genomic single disease cohort in the world; Project MinE. Dr Al Khleifat is the leading the analysis for the structural variation genome wide association analysis using 10,500 samples from the Project MinE initiative. Ammar Al-Chalabi is Professor of Neurology and Complex Disease Genetics at King’s College London, and Consultant Neurologist at King’s College Hospital, London, where he is Director of the King’s Motor Nerve Clinic. His clinical and research focus is amyotrophic lateral sclerosis (ALS). His team has developed a clinical staging system for ALS, and with others, a fundamental mechanistic hypothesis of ALS causation, showing it is likely a 6-step multistep process in which genetic factors may account for more than one step. He co-leads the international Project MinE consortium, sequencing more than 22,000 whole genomes, most from people with ALS, and with his team, has contributed to the identification of most known ALS genes.

## Abstract

**Objectives:** The use of synthetic data to supplement clinical trial placebo groups or for trial planning is rapidly gaining interest. However, there is not yet an established framework for generating synthetic data for these purposes. In this work we test two approaches to generating synthetic placebo arms for ALS trials with survival being the primary outcome variable.

**Methods:** For the first approach, we extracted sample subsets from the UK MND register (n = 308) using an evolutionary algorithm such that the subset baseline variables matched a target trials group, either people enrolled in LiCALS (n = 106) or people included in the PRO-ACT database (n = 171). We also applied trial specific exclusion criteria where possible or alternatively we applied a custom ‘time filter’. For the second approach, survival was predicted for LiCALS participants using the ENCALS model. Survival probabilities from each method were compared to real placebo participants using Kaplan-Meier analysis and the log rank test.

**Results:** We found that the synthetic placebo groups derived from the MND register matched the target trials outcomes very well. The ENCALS model produces synthetic placebo groups that are significantly different to the real placebo groups. However, when participants are censored at 6 month intervals, the ENCALS synthetic group matches the target group very well between 24 and 48 months, indicating a possible timeframe that this method could be utilised.

**Conclusion:** Both the register based approach and the ENCALS prognostic model generated synthetic placebo groups that matched placebo groups from historical trials. These methods need to be validated in prospective trials.

## Introduction

Randomised clinical trials are designed to ensure that observed differences in outcomes between groups are due to the treatment effect rather than unmeasured confounders. As such, they are the gold standard for assessing medical interventions [1]. In life-limiting diseases with no effective therapy and no viable proxy for the commonly used survival endpoint, this strategy raises ethical questions [2,3]. The use of a placebo means that a proportion of people will be denied a potentially life-saving treatment for the duration of the trial. Trial designs such as cross-over studies, where treatment arms are swapped after an interval, or designs in which everyone has active therapy after an interval, may mitigate the issue of withholding new treatments, but still mean that people are not taking a potential treatment for some considerable time [4,5].

One example condition where such issues are clearly seen is amyotrophic lateral sclerosis (ALS), a fatal adult-onset neurodegenerative disease with a lifetime risk of about 1 in 300 [6]. ALS primarily affects motor neurons of the cortex, brainstem, and spinal cord, leading to a progressive weakness and ultimately death due to neuromuscular respiratory failure, typically within 2-3 years of onset [7]. Currently, only a few disease-modifying drugs, for example riluzole and edaravone, are approved for ALS, but they do not have a beneficial effect sufficiently large to be noticeable to the patient [8,9]. Use of placebo in an ALS clinical trial due to randomisation will certainly lead to permanent irreversible damage or even death, as this is the natural history of the disease. However, not using a placebo greatly complicates interpretation of trial results and the detection of adverse events so that life-saving treatments may not be licensed.

One alternative to recruiting people into a placebo arm would be to use synthetic controls, obtained using historical control data from previous clinical trials [10] or real-world data from a disease register [11]. Alternatively, a validated prognostic model could be used to predict disease outcome in people recruited to a trial giving an expected disease trajectory (assuming no treatment) that can be compared to the real trajectory where the experimental treatment was administered [12].

These approaches have value, but also potential drawbacks. Comparison of historical control data, from previous trials or registers could be biased by improvements in patient care over time and other non-measured confounders [13]. ALS trials usually recruit from specialist centres, where the patients tend to be younger and are more likely to be male than in the general ALS population [14,15,16]. Furthermore, the delay in referral and potential delay before screening and recruitment into trials means that the cohort recruited is likely to have longer survival than the population average, since the faster progressing patients will have died or become too ill to attend [17]. A related problem is that for a condition like ALS, where there may not be many trials available, the balance between prevalent and incident patients also plays a part in the bias: a pool of patients waiting for trials to become available builds up, but as time goes on, becomes enriched for longer survivors, which is not the case when there are multiple trials recruiting and newly diagnosed patients can be recruited. These differences between the general ALS population and the population recruited to trials means that clinical databases used to derive expected rates of recruitment, event calculations, and power analyses may well lead to inaccurate conclusions.

Understanding and accurately modelling these biases would allow more accurate trial design, and dramatically reduce or possibly even abolish the need for a placebo arm. We therefore explored two strategies for such modelling, using ALS as an exemplar. The first uses real-world data to model those taking placebo in a trial, and the second uses a prognostic model.

## Methods

### Data

Population level clinical data were extracted from the MND Register for England, Wales and Northern Ireland [18]. This MND Register is a population register set up in 2015. Data are collected by specialist tertiary centres, general neurology clinics, specialist nurses and palliative care services and sent to a central repository for data cleaning and analysis. Data on people diagnosed between 01/01/2017 and 31/12/2018 were extracted from the database.

Clinical trials data were obtained from the following sources: the Lithium Carbonate in amyotrophic lateral sclerosis trial (LiCALS) [19] and the Pooled Resource Open-Access ALS Clinical Trials (PROACT) database [20]. The LiCALS clinical trial was a phase 3, randomised, double blind, placebo-controlled trial of lithium in ALS, completed in 2012, and includes 107 people randomized to lithium therapy, and 107 randomized to placebo. Survival was the primary endpoint. The PROACT database was established in 2014. It contains clinical data from 16 trials from 1990 to 2010. The use of PROACT data as a historical control group was suggested at its initial publication and it has indeed been used in this way [21]. PROACT contains data for 8635 people with ALS, including treatment/placebo grouping, survival, ALSFRS, adverse events and demographics.

We modelled the following variables: age of onset, age of diagnosis, gender, site of onset, dead or alive status at study end, survival (or time to censor date), El Escorial category, total score on the ALS Functional Rating Scale-Revised (ALSFRS-R), the presence of frontotemporal dementia, and genotype status for the C9orf72 expansion mutation.

### Approach 1 - Population register and historical control filtering and matching

The combination of biased recruitment from specialist ALS clinics and stringent trial inclusion criteria means that trial participants have different clinical characteristics from the general ALS population. Additionally, the likelihood that a patient satisfies trial inclusion criteria is tightly linked to disease progression, as trials may not be able to accommodate patients at late stages of the disease.

We developed two filter methods to mimic trial recruitment and applied them separately to the MND register data. We then used an evolutionary algorithm (EA) to select a subgroup from the filtered sets whose average baseline variables matched a real, target placebo group. We also applied the EA to historical control data from PROACT, to test whether EA baseline matching can increase the similarity of a historical and more recent control group (LiCALS). We compared the survival of the resulting synthetic placebo groups with that of the real placebo groups using the Kaplan-Meier product limit distribution and the log rank test.

#### Filter 1 – Stepwise Filter

We filtered the population register to select only those who attended a specialist clinic and then applied trial selection criteria for LiCALS: age of onset 18 to 85 inclusive and disease duration from 6 months to 3 years. Sample size changes as the filters are applied, and so to account for this at the comparison stage, data were subset to match the smallest dataset. These datasets were subset multiple times and the mean results reported with standard deviation (SD).

#### Filter 2 – Time filter

A prevalent cohort of people living with ALS will have a median survival that increases with time since diagnosis, and will deviate from survival in an incident population. The reason for this is that those still alive at any arbitrary time since diagnosis are more likely to be those without poor prognostic factors. To simulate this effect, we assigned a random number representing diagnosis date to each patient in the register with a known disease duration. We used diagnostic delay and known survival duration to calculate the week in which a patient would die:

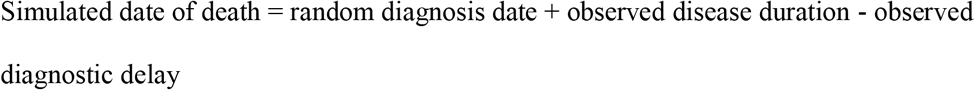

At each week, we calculated the median survival duration of register participants who had been diagnosed but had not yet died. This simulation was run 20 times to give weekly averages and the results were compared to the total MND register median, which aggregates both living and dead participants over time. We expected that the weekly median survival rate would deviate from the total register median survival with time because of the proportions of prevalent and incident patients; prevalent patients are selected for longer survival because those with short survival are removed from the pool.

Only those patients alive at a trial’s recruitment date are eligible for selection for the trial, and this contributes to preferential recruitment of long survivors into trials. We therefore adapted the simulation to achieve biased recruitment of longer surviving patients on to a trial. For every week that a patient was diagnosed but had not yet passed a certain percentage of their disease progression (60% LiCALS, 50% PROACT), they had a 5% chance of being recruited into the synthetic placebo group. The recruitment chance and percentage progression parameters were obtained using exhaustive grid search. This probabilistic time-based approach was then used as an alternative to the stepwise filter method.

#### Evolutionary algorithm

A custom, evolutionary algorithm was applied to subgroups derived from either the stepwise or time filters to extract a group whose baseline variables matched the baseline variables of a targeted trial placebo group, remaining unblinded to treatment arm as would be the case in reality. We hypothesised that matching the distribution of baseline variables in the synthetic placebo group to the real, target group, would give a better match in the outcome variable.

The EA starts with a population of randomly generated ‘agents’. These agents are ascribed a fitness level and the fittest agents ‘reproduce’ - to give a new generation. This is repeated over many iterations and the fitness of the population increases. This method can be used for numerical optimisation, where the fitness function is a function we want to maximise (or minimise) and the agents are possible solutions.

An agent in our EA was a bit vector (a list of 1s and 0s) which indexes an input dataset, giving a subset. The fitness function is minimised by the EA and was defined as a linear sum of differences in the baseline variables. Fit bit vector pairs were mated by splitting them at multiple random positions and taking alternate sections from both parents to build up a new bit vector offspring (Supplementary methods 1). As the algorithm iterates, fit agents (data subsets) whose baseline variables match the target become over-represented and we are left with a final population of data subsets whose baseline variables match the target very well. A more detailed breakdown of the algorithm is available in supplementary methods 1. Code for this EA can be found at: https://github.com/harrybPHDcode/ALS_synthetic_trial_data

The EA was applied to population data from the MND Register that had either undergone stepwise filtering or time filtering (Filters 1 and 2 above) with LiCALS means and proportions being the target. The EA was also applied to data from the PROACT clinical trials database to match placebo data from LiCALS. We called this approach ‘historical EA’.

The evolutionary algorithm is stochastic, meaning the output differs based on the random generation of the initial population and the random recombination occurring in the reproduction phase. To account for the resulting variability, the EA was run 20 times and results concatenated together to give an average result. Variance between runs was reported. The EA output was the synthetic placebo group, whose survival was then compared to the real, target group survival using the log rank test.

### Approach 2) Predicting survival of trial participants using a prognostic model

We used the baseline characteristics of trial participants to predict their survival at the trial endpoint using the ENCALS Prognostic Model, a probabilistic model of ALS survival that was developed and validated using population data on more than 11,000 people with ALS in Europe [22].

The ENCALS model requires age at motor symptom onset, months between onset to diagnosis, rate of ALSFRS-R point decline per month, site of onset of motor symptoms, El Escorial category at diagnosis, forced vital capacity at diagnosis, presence of frontotemporal dementia and whether someone carries a C9orf72 expansion mutation. It was assumed that people enrolled in trials did not have frank frontotemporal dementia as ALS trials usually exclude people with this condition. Additionally, where people did not have C9orf72 expansion mutation status recorded, this was imputed using a binary distribution with a probability of 12% likelihood of having a C9orf72 mutation, based on frequencies of people with C9orf72 variants in familial and sporadic ALS populations similar to those used for modelling.

The ENCALS model produces a set of individual survival probabilities corresponding to months from onset. To calculate survival probabilities at the group level using Kaplan-Meier analysis, for each individual we extracted a predicted survival time that corresponded with a survival probability randomly picked from a normal distribution with a mean of 0.5. In order to simulate data as it would occur throughout the course of a trial, values were truncated at different cut-off points. For example, to simulate a 12-month follow-up, observed or predicted survival greater than 12 months from trial inclusion would be categorised as censored at 12 months. For each cut off point, Kaplan-Meier survival probabilities and the log rank test were calculated 200 times. The survival probabilities and standard error from each imputation were combined by Rubin’s rules after complementary log-log transformation. The combined survival probabilities and confidence intervals were visualised compared to the observed survival using Kaplan-Meier plots.

Log rank tests were combined using a method for combination of multiply imputed Chi-squared tests. Data were analysed in R version 4.0.2 using the packages ‘survival’, ‘miceadds’ and ‘ggplot2’.

## Results

### Stepwise filter EA and historical EA

Use of stepwise filters with the population register increases cohort survival time and decreases the percentage of people with bulbar onset ALS. However, stepwise filters have no effect on sex balance, mean age of onset or mean age of diagnosis. After applying the evolutionary algorithm (EA) to this subgroup, the sex ratio was matched with those in the target dataset, and the mean ages of onset and diagnosis dropped to a value closer to that seen in the LiCALS trial. The average results are shown in Table 1 and Figure 1.

**Table 1.**
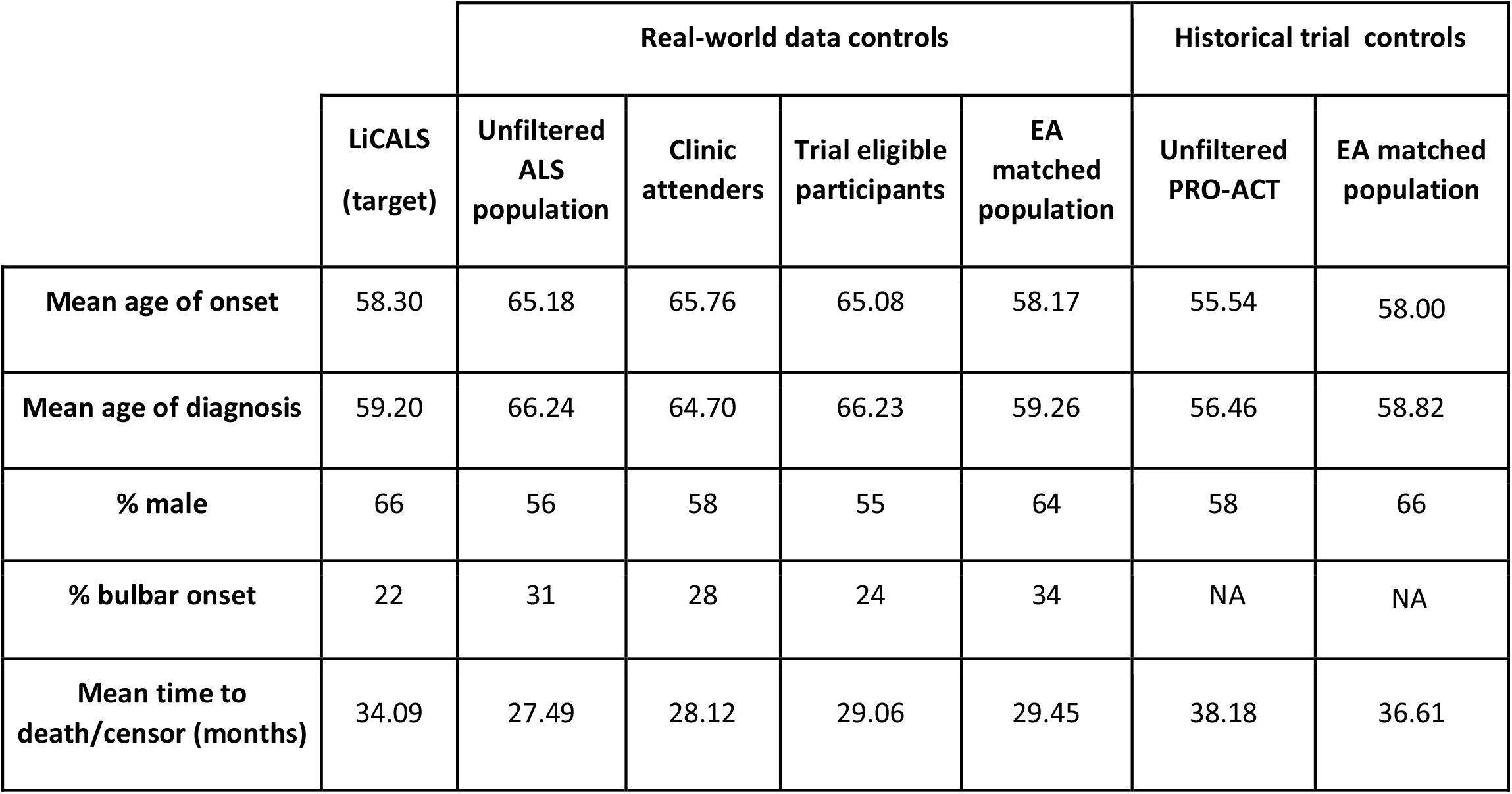
Clinical variables for the MND register (starting population), Proact (starting population) and LiCALS (target population). This table contains results for the stepwise and historical evolutionary algorithm method for generating virtual placebo populations. For the stepwise EA, we start with population register and each step of the stepwise EA filter method is represented as a column. ‘Clinic attenders’ is the first filter step, followed by ‘Selected clinic attenders’ and finally ‘Evolutionary algorithm (stepwise)’. For the historical EA method, PROACT is the starting population and the output is ‘Evolutionary algorithm (historical)’. In both cases, the LiCALS population is the target dataset.

**Table 2.** Results of Kaplan-Meier analysis of simulated vs observed populations. The mean test statistic of log rank tests and standard deviations comparing the data subsets from each step of the stepwise EA filtering with the target (LiCALS).

| | Mean test stat ± SD |
| --- | --- |
| Population register | 2.02 ± 1.39 |
| Clinic attenders | 0.71 ± 0.66 |
| Selected clinic attenders | 0.36 |
| Evolutionary algorithm | 0.28 ± 0.33 |

**Figure 1.**
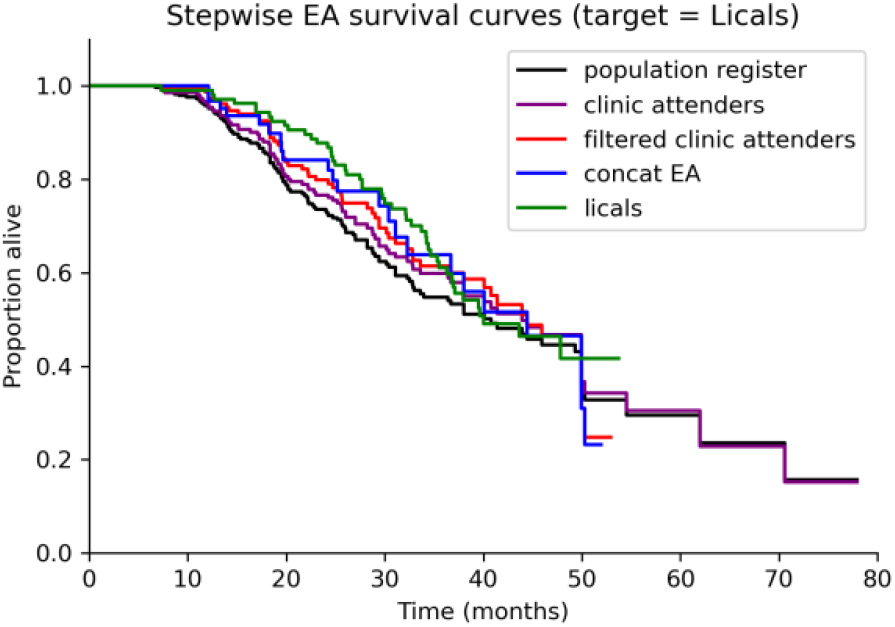
Kaplan Meier plot of Stepwise EA applied to LiCALS data. Data subsets for each step of the stepwise EA algorithm are represented as a different colour survival curve. The starting population is the black curve, the target population is the green curve. Confidence intervals have been removed to improve readability. Log rank test results for each step are reported in Table 2.

The survival curves for each step are plotted in Figure 1. Each step of the stepwise selection shifts the survival curve to the right towards the clinical trial population survival curves. Log rank tests were applied to compare survival at each step of the filtering process to the LiCALS target (Table 1). The test statistic decreases over the course of the stepwise selection process, showing that the difference between the synthetic placebo population variables and the target population variables reduces at each step.

The EA was applied to the PROACT data to select a subset whose baseline variables better matched the LiCALS group (Table 1). For example, the PROACT mean age of onset is 55.54 while for LiCALS it is 58.30. After application of the EA, the average resulting subset had a mean age of onset of 58.00. The survival of the twenty concatenated EA subsets matched the observed LiCALS placebo group survival very well (Figure 2).

**Figure 2.**
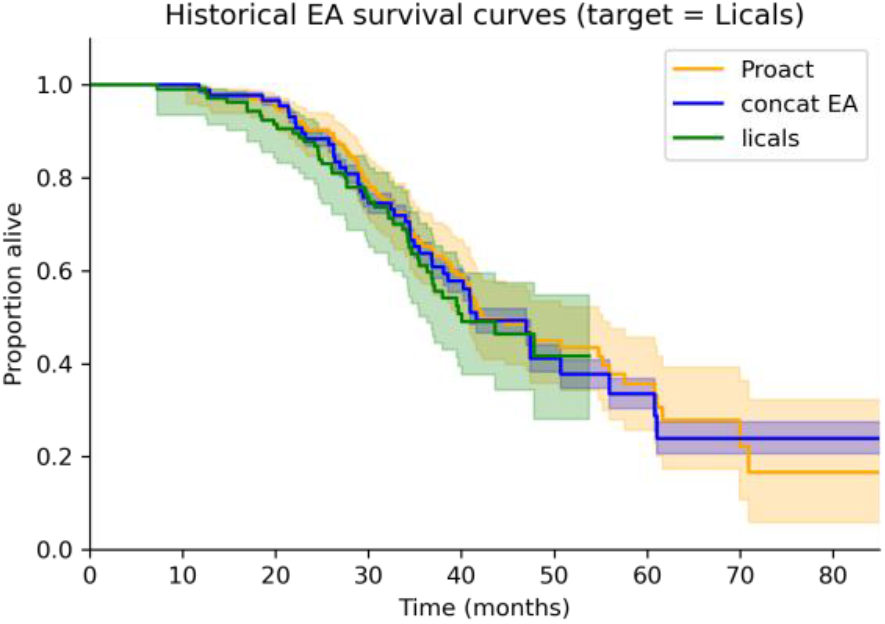
Kaplan Meier survival curve for PRO-ACT data, LiCALS data (the target population) and the simulated data using the EA. The survival curves of PROACT (orange, the starting population), concat EA (blue, concatenated results from the EA) and Licals (green, target population). The log rank test statistic for PROACT and LiCALS is 0.6. The log rank test statistic for the EA result and LiCALS is 0.34.

### MND register over time

A simulation method was developed to test the hypothesis that the aggregated data stored in the MND register reflects ALS patient demographics dependent on the mix of prevalent and incident patients in the register. Figure 3 shows how the sample survival duration changes over time (measured in weeks) as people with ALS join the register and die. Over time, the median survival duration of ALS patients deviates from the total MND register in the direction of the clinical trials placebo group medians, and this is very sensitive to the proportion of prevalent patients compared with incident patients. The median survival of the total register is higher than the median for the subset who have died, and as shown in Figure 3, increases with time more rapidly when the prevalent population is high compared with the incident population.

**Figure 3.**
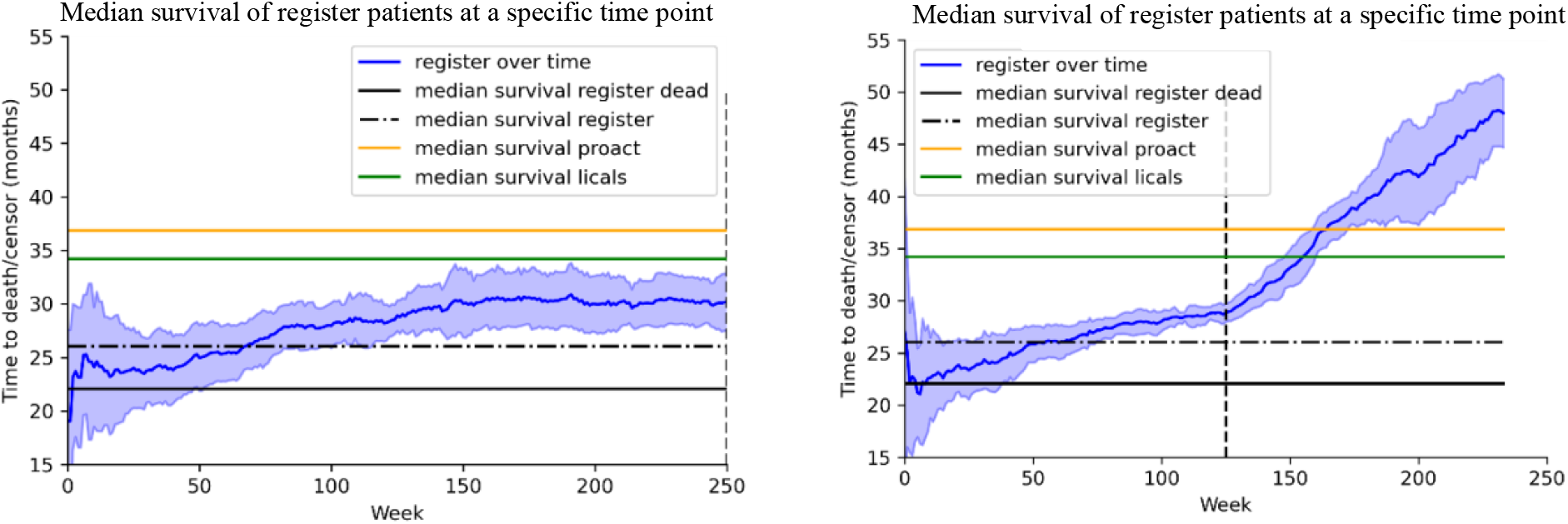
Changes in register median survival as a function of register age. The blue line is the median survival duration for the sample of people with ALS who are currently in the register and are still alive, with standard deviation given by the shaded areas. The vertical, dashed black line marks the last week that a person joins the register. Left: one new patient joins the register per week. Right: two new patient joins the register every week.

### Time-based filter EA

We repurposed the time simulation to preferentially select participants from the register with longer survival (time filter) as is seen in trial populations when a clinic has not opened a trial for some time and there is a waiting list of patients wanting to participate. The resulting subset can then be passed through the EA to give a final virtual placebo population. This method selects virtual placebo populations that are very similar to real trials populations in terms of survival (figure 4), but is highly dependent on the disease-duration cut-off, showing the importance of modelling waiting times when designing trials.

**Figure 4.**
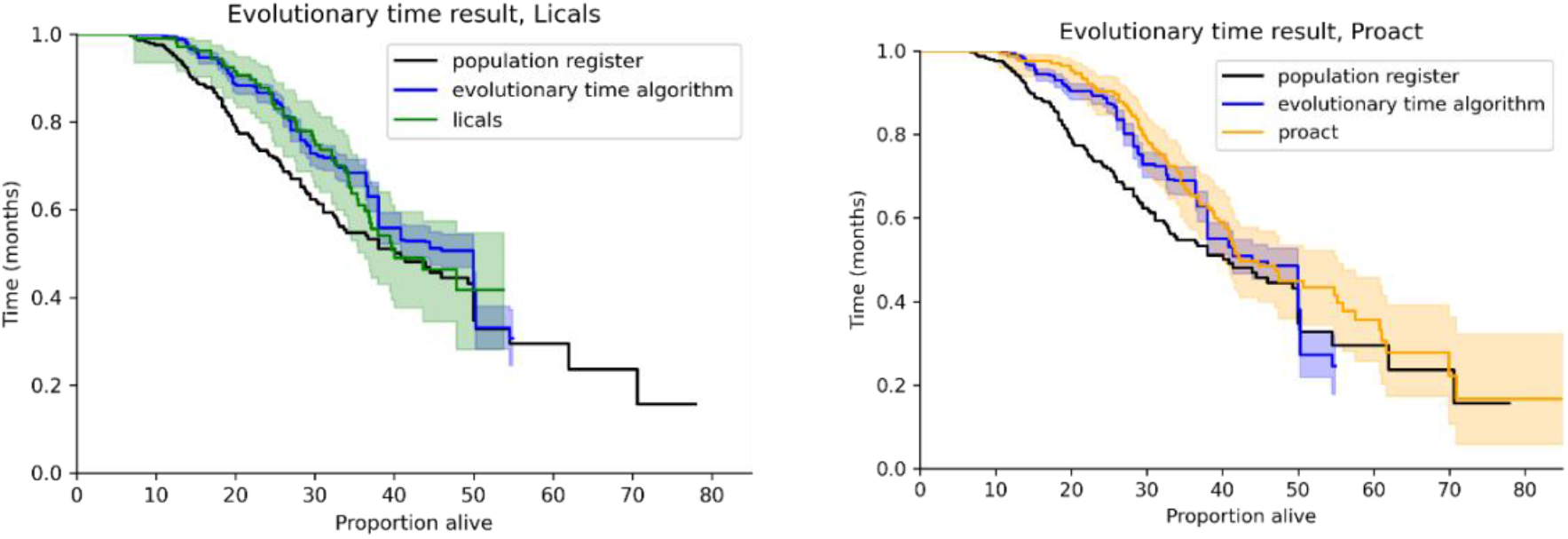
Survival curves given by the evolutionary time algorithm. Both panels show the target placebo group, the original register group and the synthetic placebo population. The left panel shows the results of the EA-time algorithm targeted to LiCALS, the right shows the EA-time result targeted to PROACT.

### Approach 2 - Prognostic model to predict survival of people in the treatment arm

Using the ENCALS model to predict survival of people in the control arm of clinical trials provides comparable estimates for survival between 24 and 48 months from onset, as shown by log rank p-values of greater than 0.05 at these time periods (Table 3). While predicted survival overall is worse than observed survival (log rank p-value <0.001), the predicted survival matches observed populations more closely when the survival times are truncated, probably due to censoring longer survivors (Figure 5).

**Table 3.** Effects of data input truncation on mismatch of observed and predicted survival for LiCALS data. This table shows p-values from log rank tests comparing the truncated predicted results to the observed LiCALS results, censoring at different time points. The optimal time points are those closely matching trial durations in the LiCALS and PROACT datasets.

| Time(months) | 12 | 18 | 24 | 30 | 36 | 42 | 48 | 54 | 60 |
| --- | --- | --- | --- | --- | --- | --- | --- | --- | --- |
| P value | 0.045 | 0.012 | 0.718 | 0.738 | 0.717 | 0.422 | 0.113 | 0.043 | 0.025 |

**Figure 5.**
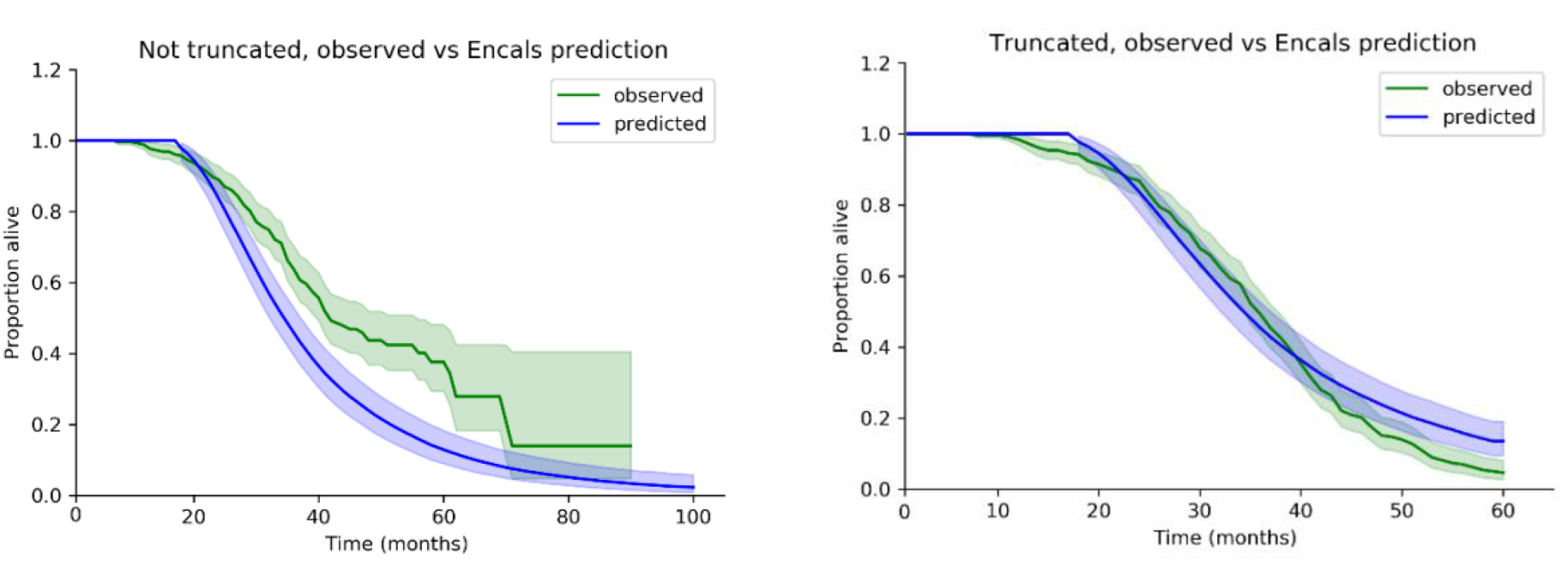
Kaplan-Meier survival curves of predicted vs observed survival using the ENCALS prognostic model. Left panel: LiCALS clinical trial, right panel: truncated input dataset, which gives closer match to real clinical trials data.

## Discussion

In this study we have shown that there are differences in clinical features between people enrolled in ALS trials and those recorded in population databases. If synthetic placebo populations are to be used in trials, a method of accounting for these many biases is required. We have shown that it is possible to use filters and an evolutionary algorithm to match the demographics of people in a dataset collected at a population level to people in the active arm of a trial. Additionally, we have found that using a prognostic model to predict survival generates comparable estimates to people in the active arm of trials, within a two-year time-frame and one year after disease onset. Synthetic placebo data could be used at the start of a trial, given baseline parameters for the recruited participants we could select a matching subset using the EA to estimate trial duration or for power analysis. At the end of a trial, with unblinded data, it would be possible to supplement the placebo group with synthetic data.

The virtual placebo population selected by the evolutionary algorithm when applied to the PROACT data matched the newer LiCALS population slightly better than the raw PROACT population. The PROACT and LiCALS survival curves were already similar, so the EA may be redundant in this specific case, but it is encouraging that the EA improved the match, and future trials depending on historical controls will need to correct for inherent differences in populations over time.

By simulating the median survival duration of register participants over time, we observe that the median survival of participants at a given time point is higher than the median of the total register, which is aggregated over time. We also observe a steep increase in median survival at the point when new participants stop being recruited to the register. This shows that, after having selected out a group of people with ALS, their median survival increases rapidly as the faster progressors die. This contributes to the over-representation of long-term survivors in ALS trials and needs to be accounted for in tools which generate synthetic trial data based on real world data. This change in median survival also impacts trial modelling, since the exact mix of prevalent and incident patients waiting for trial recruitment will not necessarily behave as expected from modelling in a population or clinic register.

ALS trials recruit from both incident and prevalent cohorts in the sense that, as fast progressors are removed from the pool of potential participants, prevalent patients are favoured. Trial exclusion criteria also prevent people very far into their disease being recruited, tending towards an incident population. We were able to recreate this mixture of incident and prevalent ALS patients by preferentially selecting long-term survivors from the MND register, mimicking the bias in real trial recruitment. The evolutionary algorithm can be applied to match the baseline variables of a target clinical trials group and select a subgroup whose demographic characteristics and survival curves match. A disease progression cut-off is needed for effective matching, because this correctly accounts for trial inclusion and exclusion criteria. For example, when modelling the LiCALS population, patients in the register more than 60% through their disease course were excluded by our algorithm. A necessary next step will be to develop a model that can automatically give the optimal cut-off parameter for a given trial, probably using factors such as trial exclusion criteria, time from recruitment to trial start, baseline participant characteristics and treatment method. There is room for improvement in the variable matching step for example, by inclusion of higher order moments or non-parametric density estimations in the fitness function or random mutations in the recombination step [23].

A limitation of this work is that we have modelled survival time, but other outcomes, including functional scores, clinical stage changes, or biomarker levels may be more informative or required as the actual trial endpoint [24]. However, survival remains an important outcome, essential for licensing of ALS drugs, and allows us to demonstrate proof of concept of the various methods explored. Another limitation is that the models are based on general ALS populations. As more trials are run on people with specific genetic variants or other subgroups, the populations used to model survival will need to be tailored to these subtypes. Another potential limitation is that the MND register itself has not been tested for recruitment bias, though we expect it to be more representative than a trial cohort, it may not be fully representative of people living with ALS in the UK.

Work on incorporating the ENCALS model into ALS trial planning and outcome has so far been focussed on characterising the prognostic profile of people with ALS enrolled in trials to refine the selection criteria and trial design [25]. As people with ALS enrolled in clinical trials tend to have a better prognosis than expected, power calculations based on survival of the general ALS population, without modifying the model as we have done, will incorrectly estimate the sample size required through overestimation of the number of expected events within the timeframe of the trial. The consequence of this is that trials need to be run for longer than expected or no difference will be shown between groups due to lack of power. For example, this observation has direct relevance to the recent Lighthouse clinical trial that used the ENCALS model to model survival [26].

We assessed the use of the ENCALS model as an alternative to a placebo arm in a trial. We found that if artificial data cut-offs are used to censor predicted survival using the ENCALS model, the survival curves match, but not if this bias is unaccounted for. Use of prognostic models to predict survival of the active arm has been investigated in cancer research [27], however the survival time in these paradigms may be much longer and is modelling remission, which is not appropriate at this stage for ALS. When there are effective treatments for ALS, the nature of parameter modelling of the placebo arm will need to be altered to take the treatment effect into account. Similarly, prognostic models should be updated periodically to reflect changing patient characteristics.

In conclusion we have shown that two different strategies can be used to create synthetic placebo populations: matching people from population registers and predicting non-treatment survival of people enrolled in trials. The results need to be replicated with new trial data, and software should be developed to aid trial design.

## Data Availability

All data produced in the present study are available upon request to the authors

## Acknowledgements

We thank people with MND and their families for their participation in this project. The authors acknowledge use of the research computing facility at King’s College London, *Rosalind* (https://rosalind.kcl.ac.uk), which is delivered in partnership with the National Institute for Health Research (NIHR) Biomedical Research Centres at South London & Maudsley and Guy’s & St. Thomas’ NHS Foundation Trusts, and part-funded by capital equipment grants from the Maudsley Charity (award 980) and Guy’s & St. Thomas’ Charity (TR130505). The views expressed are those of the author (s) and not necessarily those of the NHS, the NIHR, King’s College London, or the Department of Health and Social Care. National. The authors acknowledge Institute for Health Research (NIHR) Biomedical Research Centre at South London and Maudsley NHS Foundation Trust and King’s College London. The authors acknowledge Health Data Research UK, which is funded by the UK Medical Research Council, Engineering and Physical Sciences Research Council, Economic and Social Research Council, Department of Health and Social Care (England), Chief Scientist Office of the Scottish Government Health and Social Care Directorates, Health and Social Care Research and Development Division (Welsh Government), Public Health Agency (Northern Ireland), British Heart Foundation and Wellcome Trust. This research was supported by the National Institute for Health Research (NIHR) Biomedical Research Centre based at Guy’s and St Thomas’ NHS Foundation Trust and King’s College London. The views expressed are those of the authors and not necessarily those of the NHS, the NIHR or the Department of Health

A.I. is funded by South London and Maudsley NHS Foundation Trust; MND Scotland, Motor Neurone Disease Association, National Institute for Health Research, Spastic Paraplegia Foundation, Rosetrees Trust, Darby Rimmer MND Foundation and MND Scotland. AAK is funded by ALS Association Milton Safenowitz Research Fellowship (grant number22-PDF-609.DOI :10.52546/pc.gr.150909.), The Motor Neurone Disease Association (MNDA) Fellowship (Al Khleifat/Oct21/975-799), The Darby Rimmer Foundation, and The NIHR Maudsley Biomedical Research Centre. This project was funded by the MND Association and the Wellcome Trust. This is an EU Joint Programme-Neurodegenerative Disease Research (JPND) project. The project is supported through the following funding organisations under the aegis of JPND - www.jpnd.eu (United Kingdom, Medical Research Council (MR/L501529/1 and MR/R024804/1) and Economic and Social Research Council (ES/L008238/1)). AAC is an NIHR Senior Investigator. A.A.C. receive salary support from the National Institute for Health Research (NIHR) Dementia Biomedical Research Unit at South London and Maudsley NHS Foundation Trust and King’s College London. The views expressed are those of the authors and not necessarily those of the NHS, the NIHR or the Department of Health. The work leading up to this publication was funded by the European Community’s Health Seventh Framework Program (FP7/2007–2013; grant agreement number 259867) and Horizon 2020 Program (H2020-PHC-2014-two-stage; grant agreement number 633413). This project has received funding from the European Research Council (ERC) under the European Union’s Horizon 2020 research and innovation programme (grant agreement n° 772376 - EScORIAL.

